# Who Gets Missed? Inequities in Antenatal HIV Testing in Zambia Despite Routine Screening Policies

**DOI:** 10.1101/2025.07.10.25331268

**Authors:** Newton Nyirenda, Whiteson Mbele, Azielyn Mutiibwa, Hannah Muturi, Linda Siachalinga

**Author notes:** **Corresponding Author**: Newton Nyirenda, MD, MSc.

## Abstract

**Background:** Although substantial advances have been made in eliminating mother-to-child HIV transmission in sub-Saharan Africa, significant disparities in antenatal HIV testing by socioeconomic status and geographic location continue. This study examined predictors of HIV testing during pregnancy among women in Zambia, with a focus on equity-related factors.

**Methods:** This study utilized data from the 2007, 2013–2014, and 2018 rounds of the Zambia Demographic and Health Surveys, which are nationally representative household surveys. Eligible participants were women aged 15–49 who reported a live birth in the five years preceding the survey and received at least one antenatal care visit during that pregnancy. We applied survey-adjusted logistic regression models to analyze time trends and identify key sociodemographic factors associated with HIV testing uptake. We also conducted stratified analyses by urban–rural residence.

**Results:** HIV testing among women who attended antenatal care and had recent births rose from 87% in 2007 to 95% in 2018. However, persistent disparities were observed. Women with no education, in the poorest wealth quintile, or residing in rural areas were significantly less likely to be tested. In multivariable models, education and wealth were strong predictors of testing uptake. Stratified models revealed that education and wealth gradients were steeper in rural than urban areas.

**Conclusion:** While Zambia has made major gains in antenatal HIV testing coverage, persistent inequities remain among the poorest, least educated, and rural-dwelling women. To close these gaps, national policy should prioritize community-based testing integrated within ANC services, expand the use of mobile clinics in rural areas, and implement peer-led education initiatives targeting underserved populations. These equity-focused strategies are essential to achieving universal HIV testing in pregnancy and advancing Zambia’s PMTCT and 95–95–95 goals.

## INTRODUCTION

Human Immune Deficiency Virus (HIV) screening as part of antenatal care is an essential public health strategy for preventing mother-to-child transmission of HIV. Early identification of maternal HIV status enables timely initiation of antiretroviral therapy (ART), reducing perinatal transmission risk to less than 1% with optimal care (1,2). As a result, universal HIV screening during antenatal care (ANC) has been endorsed by the World Health Organization (WHO) as part of routine maternal health services (1). In sub-Saharan Africa, where over 90% of the world’s paediatric HIV infections occur, integrating HIV testing into ANC remains vital to controlling the epidemic (3).

Zambia has made notable progress in expanding ANC-based HIV testing as part of its prevention of mother-to-child transmission (PMTCT) strategy. According to national guidelines, all pregnant women should be tested for HIV at their first ANC visit, with re-testing offered later in pregnancy (4). Consequently, HIV testing coverage among pregnant women increased dramatically between 2007 and 2018, aided by the scale-up of provider-initiated testing and counselling (PITC), community mobilization, and ART decentralization (5). However, progress has not been uniform, and emerging evidence suggests persistent inequities in access to maternal HIV services based on socioeconomic and geographic factors (6).

Prior studies from Zambia and other low- and middle-income countries (LMICs) have documented disparities in HIV testing uptake by educational attainment, household wealth, and urban–rural residence (7–9). Women with lower education or income levels may face greater barriers to testing, including limited health literacy, structural discrimination, stigma, and logistical challenges such as transport and service availability (10–12). Even among ANC attendees, these factors may lead to missed testing opportunities and undermine the effectiveness of PMTCT programs (13,14).

Despite growing interest in addressing maternal health inequities, there is limited recent evidence on how the equity landscape of antenatal HIV testing has evolved over time in Zambia. Most demographic health surveys (DHS)-based studies have either focused on single cross-sections or broader determinants of HIV testing without stratifying by pregnancy status or ANC attendance (15). Understanding how patterns of testing among ANC users have shifted over time, and whether key equity gaps persist, can provide actionable insights for policy and implementation.

This study addresses that gap by using three rounds of nationally representative DHS data from Zambia (2007, 2013–14, and 2018) to (1) assess trends in HIV testing coverage among women with a recent birth who attended ANC, and (2) identify sociodemographic factors associated with HIV testing uptake over time. We further stratify the analysis by place of residence to examine whether observed disparities differ between rural and urban settings.

Our findings aim to inform equity-oriented strategies for achieving universal HIV testing in pregnancy, aligned with the goals of the United Nations Programme on HIV/AIDS (UNAIDS) 95–95–95 targets and the Zambia National Human Immunodeficiency Virus/Acquired Immunodeficiency Syndrome Strategic Framework.

## METHODS

### Study Design

This was a pooled cross-sectional study using data from the 2007, 2013–2014, and 2018 Zambia Demographic and Health Surveys (DHS). The DHS is a nationally representative household survey that collects information on health, fertility, maternal and child health, and HIV indicators. It uses a two-stage stratified sampling design and includes data weighted to account for complex survey design, clustering, and oversampling.

This study focused on women aged 15–49 who had a live birth in the five years preceding the survey and reported attending at least one antenatal care (ANC) visit. These inclusion criteria were used to ensure the population had reasonable opportunity to be offered HIV testing during pregnancy. For each survey, we extracted the women’s recode files (IR files) and harmonized variables across survey years to ensure comparability.

### Outcome and Explanatory Variables

The main outcome was HIV testing during pregnancy, based on self-reported receipt of an HIV test and result during the most recent pregnancy. This variable was derived from standardized DHS items collected during ANC visits (e.g., m49a_1, recoded as binary).

Key explanatory variables included survey year (2007, 2013/14, 2018), age category (<20, 20–34, ≥35), level of education (no formal education, primary, secondary, higher), household wealth index (poorest to richest), and place of residence (urban vs rural). These were constructed using DHS standard variables and selected based on prior literature highlighting their relevance to equity in maternal health service access, including studies demonstrating disparities in HIV testing uptake by age, education, wealth, and residence (16–19)

### Statistical Analysis

We adjusted for the complex sampling structure of the DHS by applying survey weights and incorporating design elements for clustering and stratification (v005, v021, and v022).

Descriptive analyses were conducted by survey year using weighted proportions to summarize sociodemographic characteristics. HIV testing prevalence and corresponding 95% confidence intervals were then estimated overall and by urban–rural residence. To identify predictors of HIV testing, we fitted multivariable logistic regression models using survey weights and quasi-binomial error structures, reporting adjusted odds ratios (aORs), 95% confidence intervals (CIs), and p-values. Separate models were run for rural and urban populations to assess potential differences in associations by residence. Analyses were conducted in R version 4.3.1 using the survey, dplyr, and gtsummary packages (20).

### Ethical Considerations

This analysis utilized anonymized secondary data from the publicly available DHS Program database. Datasets were obtained from the DHS Program website on 25th September 2024, following registration and data access approval. Ethical clearance for DHS data collection was obtained from the DHS Program and from the University of Zambia Biomedical Research Ethics Committee and the Tropical Diseases Research Centre Ethics Review Committee, which were responsible for reviewing protocols at the time of each survey. No additional ethical approval was required for this secondary analysis.

## RESULTS

Table 1 summarizes weighted background characteristics of women aged 15–49 with recent births and ANC attendance, across the 2007–2018 DHS rounds. Across all survey years, the majority of women were aged 20–34 years, with a modest increase in the proportion of adolescent mothers (<20 years) from 8.3% in 2007 to 9.8% in 2018. Educational attainment improved over time, with the proportion of women reporting secondary or higher education rising from 26% in 2007 to 41% in 2018, while those with no formal education decreased from 13% to 9.1%. Wealth distribution remained relatively stable across survey rounds. The percentage of women residing in urban areas increased slightly, from 33% in 2007 to 39% in 2018. Notably, the proportion of women who reported being tested for HIV during pregnancy increased substantially, from 87% in 2007 to 95% in 2018.

**Figure 1.**
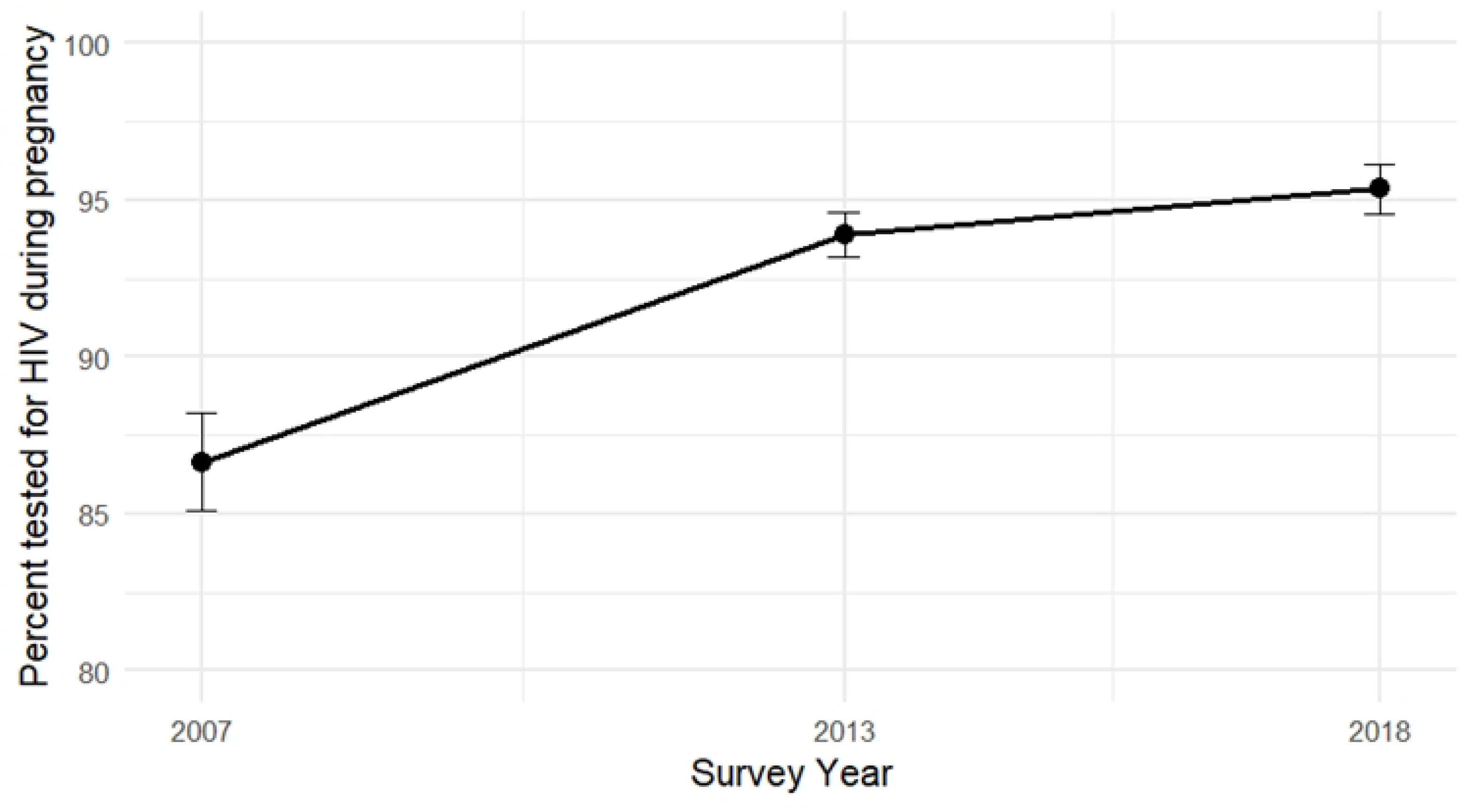
Trends in HIV testing during pregnancy among women aged 15–49 with a live birth in the five years preceding the survey and at least one antenatal care visit.

**Table 1.** Sociodemographic Characteristics of Women with a Recent Birth and Antenatal Care Attendance, Zambia DHS 2007–2018.

| Characteristic | 2007 (N = 4,047) | 2013/14 (N = 9,190) | 2018 (N = 7,245) |
| --- | --- | --- | --- |
| **Age** |  |  |  |
| <20 | 337 (8.3%) | 833 (9.1%) | 713 (9.8%) |
| 20–34 | 2,885 (71%) | 6,233 (68%) | 4,861 (67%) |
| 35+ | 825 (20%) | 2,125 (23%) | 1,671 (23%) |
| **Education** |  |  |  |

| **Characteristic** | **2007 (N = 4,047)** | **2013/14 (N = 9,190)** | **2018 (N = 7,245)** |
| --- | --- | --- | --- |
| No education | 510 (13%) | 917 (10.0%) | 662 (9.1%) |
| Primary | 2,471 (61%) | 4,919 (54%) | 3,556 (49%) |
| Secondary | 940 (23%) | 2,976 (32%) | 2,711 (37%) |
| Higher | 126 (3.1%) | 368 (4.0%) | 315 (4.4%) |
| **Wealth Quintile** |  |  |  |
| Poorest | 892 (22%) | 1,997 (22%) | 1,640 (23%) |
| Poorer | 836 (21%) | 1,932 (21%) | 1,509 (21%) |
| Middle | 818 (20%) | 1,903 (21%) | 1,379 (19%) |
| Richer | 847 (21%) | 1,775 (19%) | 1,459 (20%) |
| Richest | 654 (16%) | 1,583 (17%) | 1,258 (17%) |
| **Residence** |  |  |  |
| Rural | 2,713 (67%) | 5,693 (62%) | 4,453 (61%) |
| Urban | 1,334 (33%) | 3,497 (38%) | 2,792 (39%) |
| **HIV Tested During Pregnancy** |  |  |  |
| Yes | 3,507 (87%) | 8,623 (94%) | 6,908 (95%) |

**Table 2.** Weighted Multivariable Logistic Regression Predicting HIV Testing During Pregnancy, Zambia DHS 2007–2018.

| Characteristic | OR | 95% CI | p-value |
| --- | --- | --- | --- |
| **Survey Year** |  |  |  |
| 2007 (Ref) | — | — | — |
| 2013 | 2.3 | 1.9, 2.8 | <0.001 |
| 2018 | 3.0 | 2.4, 3.8 | <0.001 |
| **Age Group** |  |  |  |
| <20 (Ref) | — | — | — |
| 20–34 | 1.5 | 1.2, 1.8 | <0.001 |
| 35+ | 1.4 | 1.1, 1.9 | 0.008 |
| **Education Level** |  |  |  |
| No education (Ref) | — | — | — |
| Primary | 1.6 | 1.3, 1.9 | <0.001 |
| Secondary | 2.2 | 1.8, 2.8 | <0.001 |
| Higher | 1.8 | 1.1, 3.0 | 0.020 |
| **Wealth Quintile** |  |  |  |
| Poorest (Ref) | — | — | — |
| Poorer | 1.4 | 1.2, 1.7 | <0.001 |
| Middle | 1.7 | 1.4, 2.1 | <0.001 |
| Richer | 1.5 | 1.2, 1.9 | 0.001 |
| Richest | 1.4 | 0.99, 2.0 | 0.061 |
| **Place of Residence** |  |  |  |
| Rural (Ref) | — | — | — |
| Urban | 1.1 | 0.91, 1.4 | 0.300 |

\*OR = Odds Ratio; CI = Confidence Interval; Ref = Reference category.

Multivariable logistic regression analysis indicated that HIV testing uptake during pregnancy increased significantly over time. Compared to 2007, the odds of being tested for HIV were more than twice as high in 2013 (OR = 2.3, 95% CI: 1.9–2.8) and three times as high in 2018 (OR = 3.0, 95% CI: 2.4–3.8). Maternal age was also associated with HIV testing: women aged 20–34 and those 35 years or older had significantly higher odds of testing compared to those under 20 years of age (OR = 1.5 and 1.4, respectively).

Educational attainment showed a strong, positive gradient, with increasing levels of education associated with higher odds of HIV testing. For instance, women with secondary education had over twice the odds of being tested compared to those with no formal education (OR = 2.2, 95% CI: 1.8–2.8). Household wealth was positively linked with HIV testing uptake. Women belonging to the middle and higher wealth quintiles were significantly more likely to be tested during pregnancy than those from the lowest quintile. However, the association for the richest quintile was borderline significant (OR = 1.4, 95% CI: 0.99–2.0; p = 0.061). Place of residence was not significantly associated with HIV testing after adjusting for other factors (OR = 1.1, 95% CI: 0.91–1.4).

**Figure 2.**
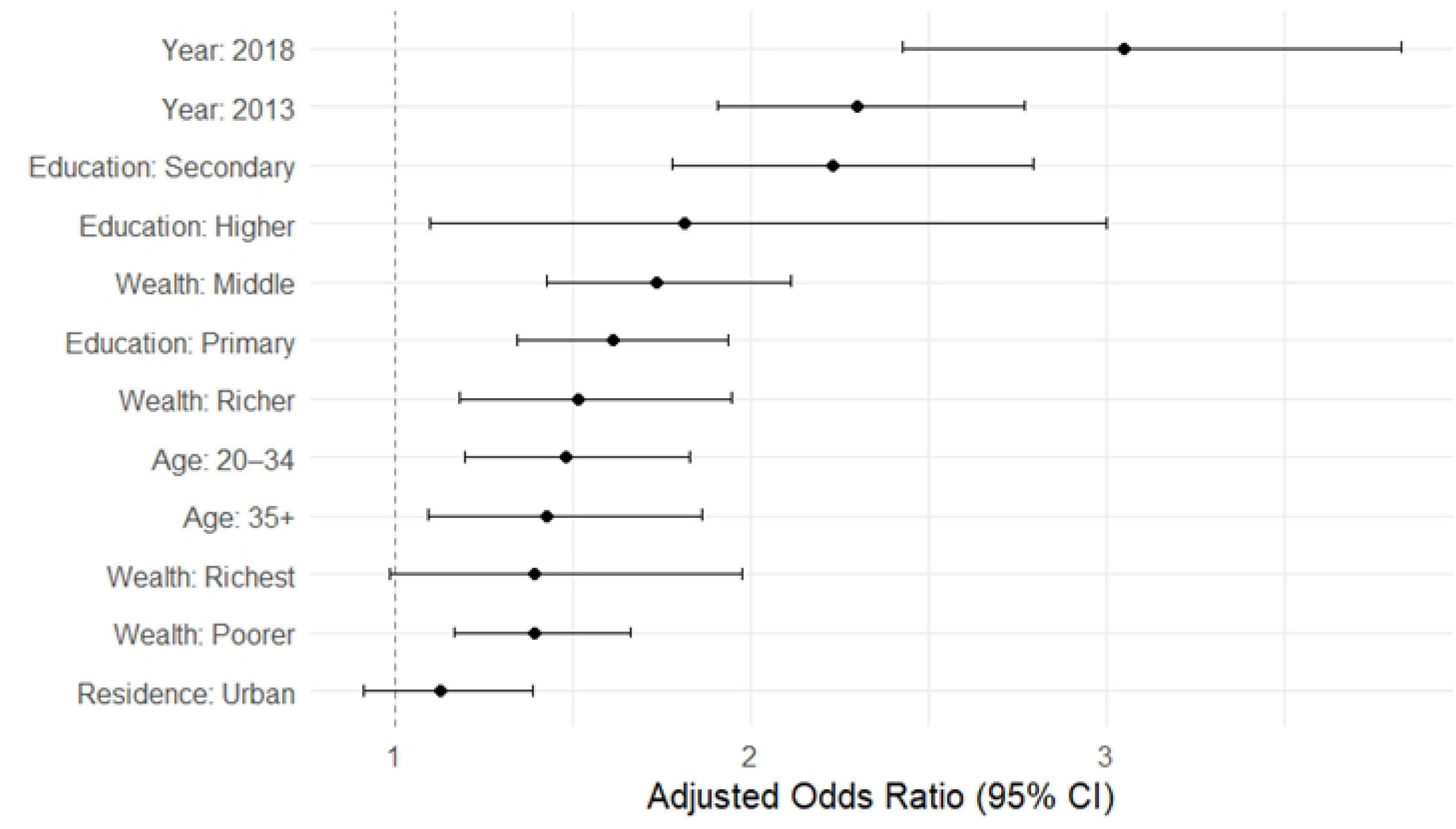
Adjusted odds ratios and 95% confidence intervals for HIV testing during pregnancy among women with a recent birth who attended antenatal care.

Stratified models revealed key differences in predictors of HIV testing during pregnancy between rural and urban areas (Table 3). In rural settings, higher education and greater household wealth were strongly associated with increased odds of testing. Women with secondary education had more than twice the odds of HIV testing compared to those with no education, and the effect of wealth was most pronounced in the richer quintiles. In contrast, these associations were attenuated and not statistically significant in urban areas. Education level was not a significant predictor of HIV testing among urban women, and only those in the poorer and middle wealth quintiles had elevated odds of testing compared to the poorest. Age-related differences were also more pronounced in rural areas, where older women were significantly more likely to be tested than adolescents, whereas no significant age effects were observed in urban areas. The trend of increased HIV testing over time was evident in both settings, but the magnitude of change was greater in rural areas, suggesting improved outreach or policy reach in underserved populations.

**Table 3.** Stratified Multivariable Logistic Regression Predicting HIV Testing During Pregnancy by Place of Residence, Zambia DHS 2007–2018.

| Characteristic | Rural Areas OR (95% CI) | p-value | Urban Areas OR (95% CI) | p-value |
| --- | --- | --- | --- | --- |
| **Survey Year** |  |  |  |  |
| 2007 (Ref) | — | — | — | — |
| 2013 | 2.10 (1.69, 2.61) | <0.001 | 2.82 (1.94, 4.09) | <0.001 |
| 2018 | 3.64 (2.86, 4.64) | <0.001 | 2.14 (1.37, 3.32) | <0.001 |
| **Age Group** |  |  |  |  |
| <20 (Ref) | — | — | — | — |
| 20–34 | 1.75 (1.38, 2.21) | <0.001 | 0.93 (0.58, 1.50) | 0.80 |
| 35+ | 1.74 (1.31, 2.31) | <0.001 | 0.81 (0.44, 1.50) | 0.50 |
| **Education Level** |  |  |  |  |
| No education (Ref) | — | — | — | — |
| Primary | 1.72 (1.42, 2.09) | <0.001 | 0.87 (0.46, 1.63) | 0.70 |
| Secondary | 2.65 (2.02, 3.47) | <0.001 | 1.10 (0.59, 2.04) | 0.80 |
| Higher | 2.24 (0.73, 6.86) | 0.20 | 0.95 (0.43, 2.13) | >0.90 |
| **Wealth Quintile** |  |  |  |  |
| Poorest (Ref) | — | — | — | — |
| Poorer | 1.37 (1.14, 1.64) | <0.001 | 2.64 (1.09, 6.40) | 0.031 |
| Middle | 1.58 (1.30, 1.93) | <0.001 | 3.65 (1.51, 8.79) | 0.004 |
| Richer | 1.95 (1.43, 2.67) | <0.001 | 2.12 (0.88, 5.08) | 0.093 |
| Richest | 1.42 (0.65, 3.10) | 0.40 | 2.22 (0.90, 5.51) | 0.085 |

\*OR = Odds Ratio; CI = Confidence Interval; Ref = Reference category.

**Figure 3.**
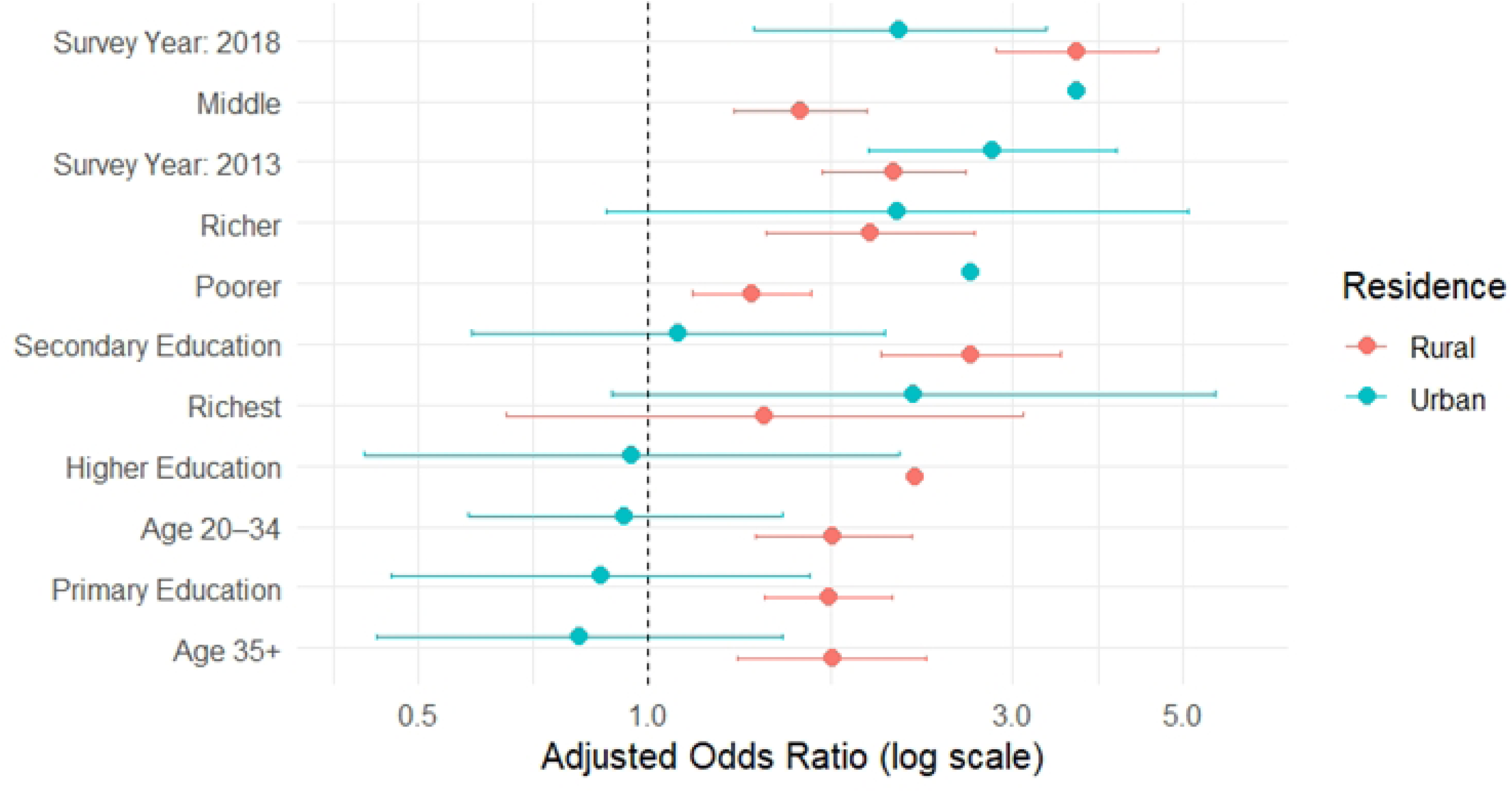
Stratified adjusted odds ratios (aORs) for HIV testing during pregnancy by place of residence, Zambia DHS 2007–2018.

## DISCUSSION

The analysis of Zambia DHS data from 2007 to 2018 indicates a substantial improvement in HIV testing during pregnancy, increasing from approximately 87% to over 95%. This trend aligns with pan-African patterns, where many countries have achieved near-universal antenatal HIV screening, although uneven across regions (21,22). The adoption of Option B+ in Zambia in 2012, providing lifelong ART to pregnant women, likely facilitated this advance by integrating HIV testing into antenatal services (23,24). National PMTCT initiatives also appear to have bolstered testing uptake during this period (25).

Despite these gains, significant disparities persist. Our results demonstrate a robust education gradient: women with secondary or higher schooling had over twice the odds of testing compared to those with no formal education, echoing findings from other East African DHS-based studies (26,27). Wealth-related disparities were also evident; middle and richer quintile women were more likely to test, consistent with broader regional analyses demonstrating pro-rich testing bias (22,26).

Age was another critical determinant, with older women showing higher testing rates, especially in rural areas, suggesting outreach strategies may be missing adolescents. Similar concerns have been reported across sub-Saharan Africa, where youth access remains limited (22,28).

### Rural-Urban Differences

Stratified analysis by residence revealed sharper socioeconomic disparities in rural than in urban areas. In urban settings, education and wealth had minimal predictive value, while in rural contexts both were strong predictors. This suggests that access barriers, due to infrastructure, information, or resources, are more pronounced outside urban centers. These findings mirror other studies illustrating how rural disadvantage amplifies socioeconomic gradients in health service uptake (21,22).

Our results have important equity implications: while overall testing coverage is high, gaps in rural, less educated, and poorer women remain. To support equity, Zambia could strengthen its national strategy by expanding rural outreach through community health workers and mobile testing clinics. Peer-led education programs tailored for women with limited formal schooling may also improve awareness and acceptance of antenatal HIV testing. In addition, integrating community-based testing into other routine maternal services can reduce both stigma and logistical barriers to uptake. These approaches are aligned with WHO and United Nations Children’s Fund (UNICEF) frameworks that emphasize the need for person-centered, context-specific care (1,29). Furthermore, international evidence supports the effectiveness of community-driven, education-sensitive models in closing gaps in prenatal HIV testing (26,28).

This study has several limitations. A key limitation of this study is that HIV testing was self-reported, which may introduce recall inaccuracies and social desirability bias. Second, the use of cross-sectional DHS data restricts our capacity to determine causal links between predictors and HIV testing outcomes. Third, we did not model geographic or regional clustering, which has been highlighted as a limitation in similar DHS-based national studies (2).

Despite these limitations, the study has notable strengths. It draws on three waves of nationally representative DHS data with consistent measures over time, allowing robust comparison of trends across more than a decade. The large sample size and use of appropriate survey weights enhance generalizability, and the stratified analysis by place of residence provides insight into urban–rural disparities that are often obscured in pooled estimates.

Furthermore, by focusing on women with recent births and antenatal care contact, the analysis highlights equity gaps within a group already engaged with the health system, offering clear direction for targeted interventions.

## CONCLUSION

HIV testing during pregnancy in Zambia has expanded substantially from 2007 to 2018, reflecting national progress toward PMTCT goals. However, despite high overall coverage, persistent inequities remain, particularly among women who are less educated, poorer, and residing in rural areas. These disparities are especially concerning given that all women in this analysis had already engaged with antenatal care services. To achieve equitable maternal HIV prevention, future programs must go beyond expanding access and address the structural, educational, and geographic barriers that limit universal testing uptake. Integrating community-based, education-sensitive strategies within antenatal services can accelerate progress toward more inclusive maternal health outcomes.

## Data Availability

The datasets used in this study are publicly available via the DHS Program website:

https://dhsprogram.com/data/available-datasets.cfm

## Declarations

### Ethics approval and consent to participate

This study used publicly available, de-identified secondary data from the DHS Program. Ethical clearance for DHS data collection was obtained from the DHS Program and relevant national bodies. No further ethical approval was required for this secondary analysis.

### Consent for publication

Not applicable.

### Availability of data and materials

The datasets used in this study are publicly available via the DHS Program website: https://dhsprogram.com/data/available-datasets.cfm

### Competing interests

The authors declare that they have no competing interests.

### Funding

This study received no specific grant from any funding agency in the public, commercial, or not-for-profit sectors.

### Authors’ contributions

NN and WM conceptualized the study, conducted the data analysis, and led the manuscript writing. LS provided contextual interpretation and policy framing. HM and AM contributed to literature review and manuscript revision. All authors reviewed and approved the final manuscript.

## Acknowledgements

We thank the DHS Program for access to the data.

## Notes

### Competing Interest Statement

The authors have declared no competing interest.

### Clinical Trial

N/A

